# Input design for unsupervised cross-national branded food database alignment using large language models

**DOI:** 10.64898/2026.05.23.26353945

**Authors:** Shinichi Nakagawa, Akira Yamamoto

**Affiliations:** Research Institute of Info-Communication Medicine (RinCOM), Tokyo, Japan; Faculty of Health Data Science, Juntendo University, Tokyo, Japan

**Author notes:** Corresponding author: Shinichi Nakagawa.

**Keywords:** food database alignment, large language model, prompt engineering, unsupervised evaluation, branded foods, nutritional epidemiology

## Abstract

Cross-national alignment of branded food databases is essential for international nutritional epidemiology but lacks standardized methods. Existing approaches — including food ontologies, domain-specific fine-tuned language models, and manual expert mapping — require either substantial infrastructure or do not scale to thousands of items. We propose an unsupervised evaluation framework for large language model (LLM)-based food database alignment that requires no ground-truth labels. Using the Japan Branded Food Database (JBFD; 9,519 items, 71 mid-level categories) and USDA FoodData Central (448 categories) as a case study, we introduce two complementary metrics: weighted centroid distance (nutritional proximity between matched category pairs) and dominant category share (structural consistency of category-level assignments). We then conducted a systematic ablation study across eight input conditions (A–H), varying combinations of product name, nutrient profile, and semantic category label. Results showed that nutrient-only inputs yielded poor structural consistency despite low centroid distances, while semantic category labels achieved the highest dominant category share (89.3%) but introduced circularity due to their LLM-derived origin. Among circularity-free conditions, product name combined with minimal nutrient information (energy, protein, salt; condition E) achieved the best balance of centroid distance (0.471) and dominant category share (65.8%). Model comparison across Claude Haiku, Sonnet, and Opus confirmed that NO_MATCH rates were consistent across model sizes (12–14%), suggesting that prompt design contributes more to alignment quality than model scale. These findings provide practical guidance for input design in LLM-based food database alignment without ground-truth annotation.

## 1. Introduction

International nutritional epidemiology increasingly depends on the ability to compare dietary intake across countries. Such comparisons require alignment between national food composition databases (FCDBs), which differ substantially in structure, cultural scope, and classification logic. The challenge is compounded when one system is flat and commercially oriented — such as USDA FoodData Central with 448 branded food categories — and the other is hierarchical and culturally specific, such as the Japan Branded Food Database (JBFD) with 71 mid-level categories derived from the Japanese food composition standard.

Three broad strategies have been proposed for cross-national food database alignment. First, unified food ontologies such as FoodOn consolidate food concepts across systems but require extensive expert curation and remain incomplete for commercially branded products [1]. Second, domain-specific fine-tuned language models — including FoodyLLM [2] and FoodSEM — achieve high classification accuracy but demand large annotated training corpora and specialized computational infrastructure. Third, manual mapping by nutrition experts, as employed in the PURE study [3], is accurate but inherently limited in scale. None of these approaches addresses the evaluation problem directly: how can alignment quality be assessed when no ground-truth correspondence exists between two heterogeneous classification systems?

This evaluation problem is fundamentally different from clinical coding alignment, where exact correspondence is mandated for patient safety and authoritative mapping tables exist (e.g., ICD-10 to SNOMED-CT). Food classification tolerates ambiguity — a Japanese instant noodle product may reasonably map to multiple USDA categories — making manual validation at scale impractical and motivating the need for automated, label-free evaluation frameworks.

We address this gap by proposing an unsupervised evaluation framework for LLM-based food database alignment. Using general-purpose LLMs with iterative prompt engineering — without fine-tuning or ontology development — we introduce two complementary metrics that capture different dimensions of alignment quality. We further conduct a systematic ablation study across eight input conditions to identify which combinations of product name, nutrient profile, and semantic category label best support alignment. The approach is validated on a Japan–US case study involving 9,519 branded food items.

## 2. Methods

### 2.1 Databases

The Japan Branded Food Database (JBFD v3) comprises 9,519 branded food items collected from Japanese manufacturer websites, organized into 71 mid-level categories under a three-tier hierarchy derived from the Japanese food composition standard (MEXT 8th edition) [6]. Nutritional data — energy, protein, fat, carbohydrate, and salt equivalent — were obtained directly from product labeling. The JBFD mid-level category labels were themselves derived through LLM-based classification in prior work [7,8], a fact relevant to the interpretation of conditions using these labels as input (see Section 4.3).

The USDA FoodData Central branded food dataset (October 2024 release) [5] contains 448 food categories. Sodium values were used in place of salt equivalent for USDA items (conversion: salt equivalent g = sodium mg × 2.54 / 1,000).

### 2.2 Prompt engineering iterations (v1–v4)

Four prompt versions were developed iteratively using Claude Sonnet (claude-sonnet-4-20250514; Anthropic) [11] applied to all 9,519 JBFD items in batches of 20.

**v1** (product name only): Items were presented by product name alone with a list of USDA categories and representative examples. No explicit NO_MATCH guidance was provided.

**v2** (Japan-specific food list): A curated list of Japan-specific foods (natto, miso, fish paste products, seaweed, dried seafood, rice crackers, traditional confectionery) was added with explicit instruction to assign NO_MATCH for these items.

**v3** (nutrition-first; pilot only): Nutrient profiles were given highest priority over product names. A 100-item pilot revealed systematic semantic misclassification (e.g., miso assigned to cheese-type categories due to similar sodium and protein profiles), and this condition was discontinued.

**v4** (three-tier priority): JBFD mid-level category name was given first priority, nutrient profile second, and product name third. This became the reference condition for subsequent analyses.

### 2.3 Evaluation metrics

*Weighted centroid distance* measures nutritional proximity between matched category pairs. For each JBFD mid-level category j matched to a USDA category u, the centroid of JBFD items in j and the centroid of USDA Foundation Foods items in u are computed in five-dimensional nutrient space (energy, protein, fat, carbohydrate, sodium). All values are standardized using a pooled StandardScaler. Euclidean distance between centroids is computed per matched pair, and the overall score is the item-count-weighted mean across all pairs with ≥3 items in both categories. Lower values indicate better nutritional alignment.

*Dominant category share* measures structural consistency of assignments within each JBFD mid-level category. For category j with n matched items assigned across multiple USDA categories, dominant category share is defined as the proportion assigned to the single most frequent USDA category. The overall score is the unweighted mean across all JBFD mid-level categories. Higher values indicate more consistent, interpretable alignment.

*NO_MATCH rate* is the proportion of JBFD items for which no appropriate USDA category was identified. This is treated as a valid outcome reflecting cultural specificity rather than classification failure.

### 2.4 Ablation study

To identify optimal input design, we conducted a systematic ablation across eight input conditions (Table 1) applied to a fixed random sample of 500 JBFD items (random seed = 42) using Claude Sonnet. All conditions used the same USDA category reference list with mean nutrient values per category and the same NO_MATCH eligibility rules for Japan-specific foods.

**Table 1.** Input conditions in the ablation study.

| Condition | Input features | Rationale |
| --- | --- | --- |
| A | Product name only | Semantic baseline |
| B | Nutrients: E, P, F, C, Salt | Nutrition-only baseline |
| C | Nutrients: E, P, Salt | Minimal nutrient set (PFC-equivalent) |
| D | Product name + E, P, F, C, Salt | Name + full nutrient profile |
| E | Product name + E, P, Salt | Name + minimal nutrients (recommended) |
| F | JBFD mid-level category name | Semantic label only (circularity risk) |
| G | Category name + E, P, F, C, Salt | Label + full nutrient profile (circularity risk) |
| H | Category name + product name + nutrients | Full input, v4-equivalent (circularity risk) |

### 2.5 Model comparison

To assess prompt-model interaction, the v4 prompt was applied to Claude Haiku (claude-haiku-4-5-20251001; 100 items, random seed = 42) and Claude Opus (claude-opus-4-5; 1,000 items, random seed = 123) and compared against the Sonnet full-run results on the same items.

## 3. Results

### 3.1 Prompt iteration results (v1–v4)

Table 2 summarizes the three evaluation metrics across prompt versions applied to the full JBFD dataset.

**Table 2.** Evaluation metrics across prompt versions v1–v4 (full dataset, n = 9,519).

| Version | NO_MATCH (%) | Weighted centroid distance | Dominant category share (%) |
| --- | --- | --- | --- |
| v1 (product name only) | 1.0 | 0.725 | 58.6 |
| v2 (Japan-specific list) | 18.3 | 0.652 | 62.1 |
| v3 (nutrition-first; 100-item pilot) | 3.0 | 0.628 | 74.4 |
| v3 full run (7,959 items)* | 0.7 | 0.413 | 54.6 |
| v4 (three-tier priority) | 14.3 | 0.607 | 69.5 |

\* v3 full run included for completeness; discontinued after pilot due to semantic misclassification.

Weighted centroid distance improved monotonically from v1 to v4. The NO_MATCH rate increased appropriately from v1 (1.0%, near-zero due to forced assignment) to v2 (18.3%) and stabilized at v4 (14.3%). The v3 pilot — despite showing a low centroid distance — was discontinued after systematic semantic misclassification was detected (e.g., miso → Cheese-type categories; instant noodles → Nuts/Seeds), demonstrating that distance alone is insufficient as an alignment quality metric. When v3 was run on all 9,519 items, the NO_MATCH rate was 0.7% and weighted centroid distance was 0.413 — the best distance score across all conditions — yet dominant category share collapsed to 54.6%, confirming the failure mode identified in the pilot.

**Figure 1.**
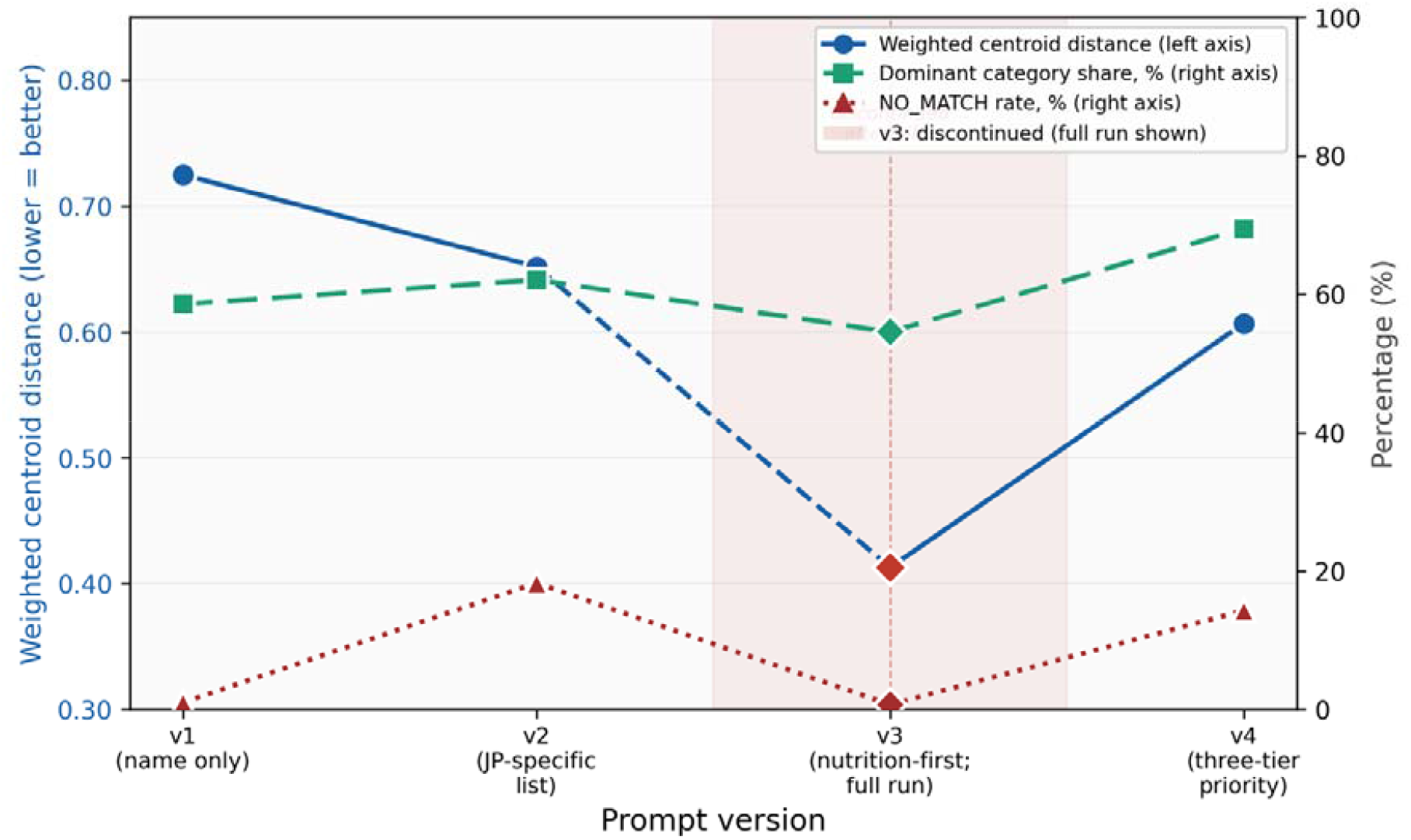
Evaluation metrics across prompt versions v1–v4. Weighted centroid distance (left axis) decreased monotonically from v1 to v4. v3 full-run showed the lowest distance but collapsed dominant category share, confirming the three-metric framework is necessary for alignment evaluation.

### 3.2 Ablation study results (conditions A–H)

Table 3 presents results for all eight input conditions. Nutrient-only conditions (B and C) showed the lowest dominant category share (48.0% and 47.2%) and lowest v4 agreement (16.2% and 13.8%), confirming that nutritional values alone are insufficient for semantically coherent alignment. Condition C (E+P+Salt) outperformed condition B (5 nutrients) on centroid distance (0.975 vs 1.403), suggesting that additional nutrient dimensions introduce noise rather than signal. Condition F (mid-level category name only) achieved the highest dominant category share (89.3%) but is subject to circularity. Among circularity-free conditions, condition E achieved the best centroid distance (0.471) with acceptable dominant category share (65.8%) and NO_MATCH rate (11.8%).

**Table 3.** Ablation study results for input conditions A–H (n = 500 per condition).

| Cond. | Input | NO_MATCH (%) | Weighted distance | Dom. share (%) | v4 agree. (%) |
| --- | --- | --- | --- | --- | --- |
| A | Product name | 12.4 | 0.373 | 65.5 | 63.6 |
| B | Nutrients (5 items) | 3.4 | 1.403 | 48.0 | 16.2 |
| C | E + P + Salt | 4.2 | 0.975 | 47.2 | 13.8 |
| D | Name + nutrients (5) | 12.0 | 0.535 | 67.7 | 66.6 |
| E† | Name + E + P + Salt | 11.8 | 0.471 | 65.8 | 66.4 |
| F‡ | Mid-level category name | 11.8 | 0.580 | 89.3 | 55.8 |
| G‡ | Category + nutrients (5) | 13.0 | 0.665 | 79.7 | 62.0 |
| H‡ | Category + name + nutrients | 16.2 | 0.641 | 78.0 | 77.2 |

† Recommended condition. ‡ Conditions F–H use LLM-derived JBFD mid-level category labels; results may reflect LLM self-consistency (see Section 4.3).

**Figure 2.**
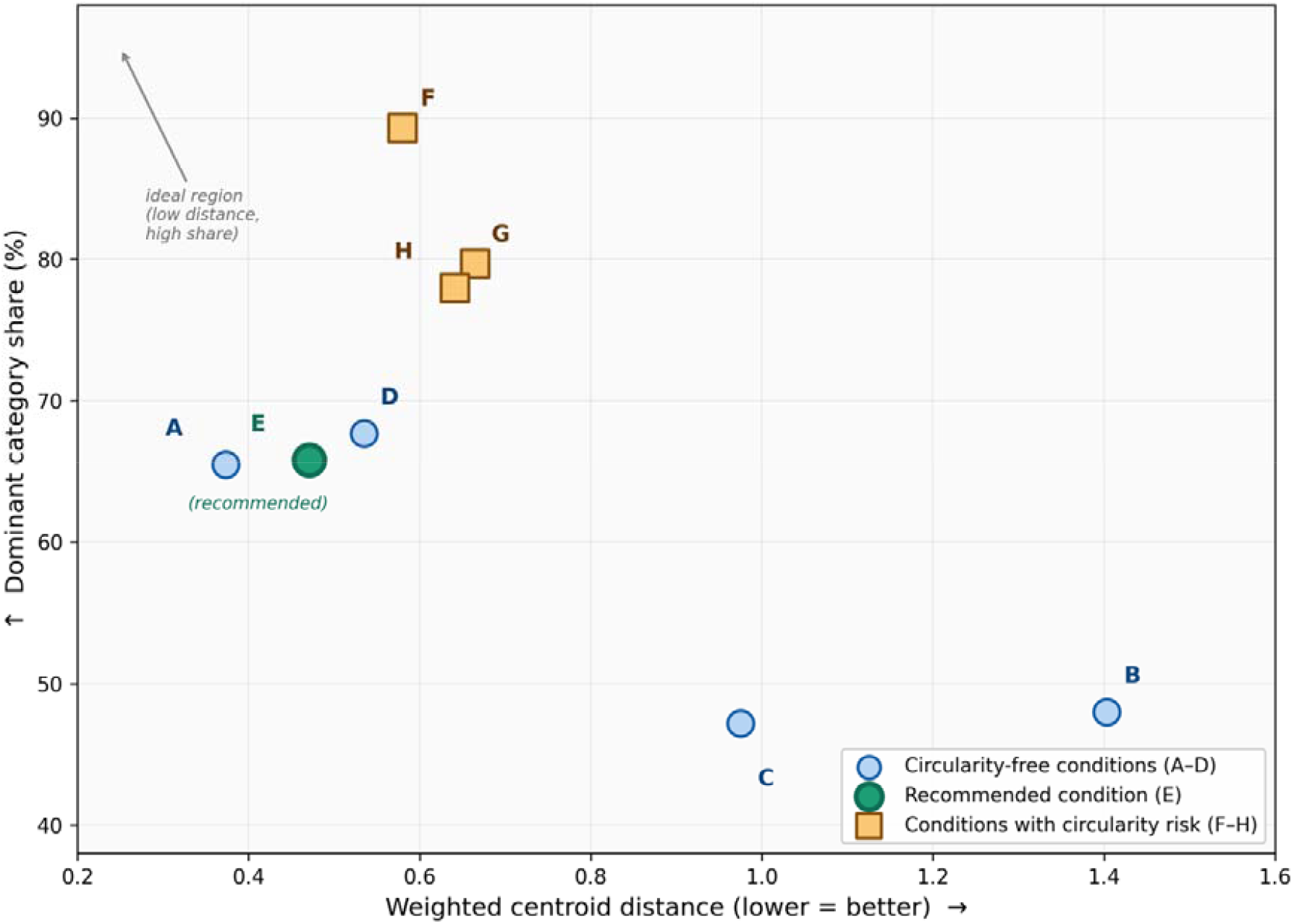
Scatter plot of weighted centroid distance versus dominant category share for conditions A–H. Circularity-free conditions (A–E, circles) and conditions with circularity risk (F–H, squares) are distinguished. The recommended condition E is highlighted. The ideal region is the upper-left quadrant.

### 3.3 Model comparison

Table 4 presents NO_MATCH rates and inter-model agreement across three model sizes. NO_MATCH rates were consistent across Haiku, Sonnet, and Opus (12–14%), and inter-model agreement with Sonnet was approximately 70% for both Haiku and Opus, despite an approximately 15-fold difference in per-token cost between Haiku and Opus.

**Table 4.** Model comparison using the v4 prompt.

| Model | Sample size | NO_MATCH (%) | Agreement with Sonnet (%) |
| --- | --- | --- | --- |
| Claude Haiku | 100 | 12.0 | 70.0 |
| Claude Sonnet (reference) | 9,519 | 14.3 | — |
| Claude Opus | 1,000 | 14.3 | 70.7 |

## 4. Discussion

### 4.1 Unsupervised evaluation as a necessary framework

Food database alignment differs from clinical coding alignment in a fundamental way: no authoritative ground truth exists. A Japanese instant noodle product may reasonably correspond to multiple USDA categories, and no regulatory mandate resolves this ambiguity. This property makes conventional supervised evaluation inapplicable at scale. The weighted centroid distance and dominant category share metrics proposed here operationalize alignment quality without ground-truth labels, measuring nutritional proximity and structural consistency respectively.

### 4.2 The failure modes of single-metric evaluation

The v3 full-run results illustrate why single-metric evaluation is insufficient. Nutrition-first prompting achieved the lowest centroid distance (0.413) across all conditions, yet dominant category share collapsed to 54.6% and semantic misclassification was pervasive (miso → Cheese; noodles → Nuts). Conversely, condition F achieved the highest dominant category share (89.3%) but cannot be used as an independent benchmark due to circularity. The two metrics together expose failure modes that neither captures alone.

### 4.3 Circularity in condition F and implications for benchmark design

JBFD mid-level category labels were derived through LLM-based classification in prior work [7,8]. Using these labels as input to the same class of model (condition F) introduces a form of self-consistency measurement rather than independent validation. The high dominant category share in condition F (89.3%) likely reflects LLM self-consistency rather than genuine semantic utility of the labels. Researchers applying this framework should treat any LLM-derived feature as a potentially circular input and design ablation conditions accordingly.

### 4.4 Practical input design recommendations

Among circularity-free conditions, condition E (product name + E+P+Salt) is recommended as the practical default. It achieves the lowest centroid distance (0.471), maintains acceptable dominant category share (65.8%) and NO_MATCH rate (11.8%), and requires only three nutrient values — often the minimum disclosed under mandatory food labeling regulations in Japan and other countries. The addition of fat and carbohydrate (condition D) modestly improved dominant category share but worsened centroid distance, consistent with the B vs C comparison showing that additional nutrient inputs may introduce noise.

### 4.5 Model scale and prompt design

The consistency of NO_MATCH rates across Haiku, Sonnet, and Opus (12–14%) and similar inter-model agreement (∼70%) suggest that the v4 prompt design is the primary driver of alignment behavior, with model scale playing a secondary role. Cost-efficient deployment with smaller models is feasible without substantial quality degradation for this task type.

### 4.6 Limitations

Several limitations warrant note. First, the JBFD covers Japanese branded foods from manufacturer websites; coverage of regional and artisanal products is limited. Second, the USDA Foundation Foods subset used for centroid calculation differs from the branded food dataset used for category reference, introducing a potential mismatch in nutritional representativeness. Third, LLM outputs are non-deterministic and model-version dependent; reproducibility requires documentation of exact prompt text, model version, and temperature settings. Fourth, the ablation study used 500 items; replication with larger samples may refine condition rankings. Fifth, generalizability to country pairs other than Japan–US has not been demonstrated, though the framework design is country-agnostic.

## 5. Conclusion

We proposed an unsupervised evaluation framework for LLM-based cross-national branded food database alignment, combining weighted centroid distance and dominant category share as complementary metrics. Applied to a Japan–US case study (9,519 items), the framework revealed that nutrition-only inputs produce nutritionally proximate but semantically incoherent alignments, while semantic category labels introduce circularity when LLM-derived. Among circularity-free input conditions, product name combined with minimal nutrient information (energy, protein, salt) achieved the best overall performance. Model scale had limited impact on alignment quality compared to prompt design. This framework lowers the barrier to cross-national nutritional epidemiology for research groups without access to annotated training data or specialized computational infrastructure.

## Data Availability

The JBFD v3 dataset and prompt texts for v1-v4 are available at [repository URL to be confirmed upon acceptance]. USDA FoodData Central data are publicly available at https://fdc.nal.usda.gov/.

## Acknowledgments

The authors thank the manufacturers whose publicly available nutritional information formed the basis of the JBFD. LLM-based classification was performed using the Claude API (Anthropic). This research received no specific grant from any funding agency in the public, commercial, or not-for-profit sectors.

## Conflict of Interest

The authors declare that they have no conflict of interest.

## Data availability

The JBFD v3 dataset and prompt texts for v1–v4 are available at [repository URL to be confirmed upon acceptance]. USDA FoodData Central data are publicly available at https://fdc.nal.usda.gov/.

## Notes

### Competing Interest Statement

The authors have declared no competing interest.

